# A SARS-CoV-2 negative antigen rapid diagnostic in RT-qPCR positive samples correlates with a low likelihood of infectious viruses in the nasopharynx

**DOI:** 10.1101/2022.03.17.22272008

**Authors:** Isadora Alonso Corrêa, Débora Souza Faffe, Rafael Mello Galliez, Cássia Cristina Alves Gonçalves, Richard Araújo Maia, Gustavo Peixoto da Silva, Filipe Romero Rebello Moreira, Diana Mariani, Mariana Freire Campos, Isabela de Carvalho Leitão, Marcos Romário de Souza, Marcela Sabino Cunha, Érica Ramos dos Santos Nascimento, Liane de Jesus Ribeiro, Thais Felix Cordeiro da Cruz, Cintia Policarpo, Luis Gonzales, Mary A Rodgers, Michael Berg, Roy Vijesurier, Gavin A Cloherty, John Hackett, Orlando da Costa Ferreira, Terezinha Marta Pereira Pinto Castiñeiras, Amilcar Tanuri, Luciana Jesus da Costa

## Abstract

SARS-CoV-2 transmission occurs even among fully vaccinated individuals; thus, prompt identification of infected patients is central to control viral circulation. Antigen rapid diagnostic tests (Ag-RDT) are highly specific, but sensitivity is variable.Discordant RT-qPCR vs Ag-RDT results are reported, raising the question of whether negative Ag-RDT in positive RT-qPCR samples could imply the absence of infectious viruses. To study the relationship between a negative Ag-RDT results with virological, molecular, and serological parameters, we selected a cross sectional and a follow-up dataset and analyzed virus culture, subgenomic RNA quantification, and sequencing to determine infectious viruses and mutations. We demonstrated that a positive SARS-CoV-2 Ag-RDT result correlates with the presence of infectious virus in nasopharyngeal samples. A decrease in sgRNA detection together with an expected increase in detectable anti-S and anti-N IgGs was verified in negative Ag-RDT / positive RT-qPCR samples. The data clearly demonstrates the less likelihood of a negative Ag-RDT sample to harbor infectious SARS-CoV-2 and consequently with a lower transmissible potential.

## Introduction

Since December 2019, more than 250 million confirmed cases and 5 million deaths have been attributed to SARS-CoV-2-related infections worldwide (*1*). Vaccines against COVID-19 became available at the end of 2020, and as of 12 November 2021, 3.1 billion people were fully vaccinated (41% of the world’s population). Although vaccination may reduce COVID-19 cases and disease severity (*2,3*), SARS-CoV-2 breakthrough infection still occurs amongst fully vaccinated subjects (*4,5,6*). In this scenario, social distancing measures and early detection/isolation of infected individuals remain central for viral dispersion control. SARS-CoV-2 diagnosis relies on the gold-standard technique of nucleic acid amplification tests (NAATs), and several assays targeting different viral genes are available and widely used (*7*). In Brazil, NAATs are recommended by Ministry of Health only for hospitalized patients with severe respiratory syndrome, healthcare workers presenting flu-like symptoms and organ donates candidates; with 3 to 15 days from the beginning of symptoms (8). However, the high costs and operational requirements limit its availability. Besides, NAATs long turnaround time to get results may negatively impact the clinical outcome and epidemiology. Thus, strategies to accelerate diagnosis are still required.

SARS-CoV-2 Ag-RDTs represent powerful tools to expand the number of people tested and increase testing frequency (*8,9,10*). Most commercial Ag-RDTs, based on lateral flow immunoassays, capture SARS-CoV-2 Nucleocapsid (N) protein due to their high degree of conservation and the proportionally high amounts in coronavirus virions (*11,12*).

Currently, WHO recommends the use of Ag-RDTs as a diagnostic tool when the tests achieve a minimum of 80% sensitivity and 97% specificity, compared to a reference NAAT. Ag-RDTs can be especially useful in locations where NAAT is unavailable or has delayed time responses (*13*). Indeed, several studies report high specificity with few variations among Ag-RDTs assays in clinical samples (*10,14,15,16,17*). Most studies also demonstrate a correlation between the number of days since symptom onset (DSSO), viral load, and sensitivity. Ag-RDTs’ sensitivity varies from 85% to 94% when used 3-5 DSSO (*14,15,18*) with high sensitivity for samples with C_t_ values below 25 (*14,18,19*).

In addition to its low cost and quick result, preliminary studies have also demonstrated a reliable correlation between a positive Ag-RDT result and viral isolation in cell culture, supporting Ag-RDTs as a convenient tool for early detection of high virus spreading individuals, enabling appropriate patient isolation and improving transmission control (*20,21*). Although its cause remains unclear, a small proportion of patients show a negative Ag-RDT result with detectable viral load by NAAT, and positive viral culture (*22*). Thus, these discordant cases (Ag-RDT+/NAAT-) could impact the spread of SARS-CoV-2 (*22,23*).

Here, we analyzed patients with positive SARS-CoV-2 RT-qPCR result and negative Ag-RDT using the Panbio™ COVID-19 Ag test (Abbott) from a cohort of patients attending for SARS-CoV-2 diagnosis at the Federal University of Rio de Janeiro, Brazil, from August 2020 to September 2021. We found that 10-35% of samples that tested positive by SARS-CoV-2 RT-qPCR were negative at Ag-RDT consistently throughout the period of the study, regardless of the predominant SARS-CoV-2 variant circulating in Rio de Janeiro State. We further characterized 23 concordant positive (RT-PCR+/Ag-RDT+) and 29 discordant (RT-PCR+/Ag-RDT-) samples for virus isolation in cell cultures, RT-qPCR correlation, presence of anti-Spike and anti-Nucleocapsid IgG, full-length viral genome and viral protein content. We found that discordant samples harbor intact full-length genomes, but only 10.53% were replication-competent compared to 56.52% of the true positive counterpart. Through statistical models and *in vitro* assays, we demonstrated that the two variables with greater predictable capacity for a positive Ag-RDT result were the RT-qPCR C_t_ value and the presence of a humoral response against SARS-CoV-2.

## Material and Methods

### Study samples

The Federal University of Rio de Janeiro offers diagnostic tests for mildly symptomatic public health care and security force workers in the city of Rio de Janeiro and to the University community. For patients presenting up to five days from symptom onset at the time of testing, a swab was taken to perform the Panbio™ COVID-19 Ag test. Additional nasopharyngeal swabs were collected from both nostrils using two rayon-tipped swabs for RT-qPCR testing. The Viral Transport Media (VTM) sample was then used for RNA extraction and RT-qPCR for SARS-CoV-2 detection, and the leftover sample was stored at −80°C for further inoculation in cell culture. All patients were requested to take another RT-qPCR test 14 days after the initial symptoms and, if remained positive, to take a follow-up test every 7 days until a negative RT-qPCR result was obtained. Plasma and serum were also collected during each visit for serological test purposes.

We selected samples from August-2020 to September-2021 in two study cohorts: a cross-sectional cohort composed of patients that, at the time of diagnosis, presented an antigen test that was further confirmed (antigen concordant) or not (antigen discordant) by RT-qPCR; and a follow-up cohort, composed of patients that, at the time of diagnosis, presented antigen and RT-qPCR positive results and had at least one more sample collected with a minimum six day interval where antigen and RT-qPCR results were concordant or discordant. For all the selected samples, 100 uL of the VTM sample was tested again with the Panbio™ COVID-19 Ag test according to the manufacturer’s instructions. A flow chart of the sample selection and the study’s cohorts is shown in Figure 1.

**Figure 1.**
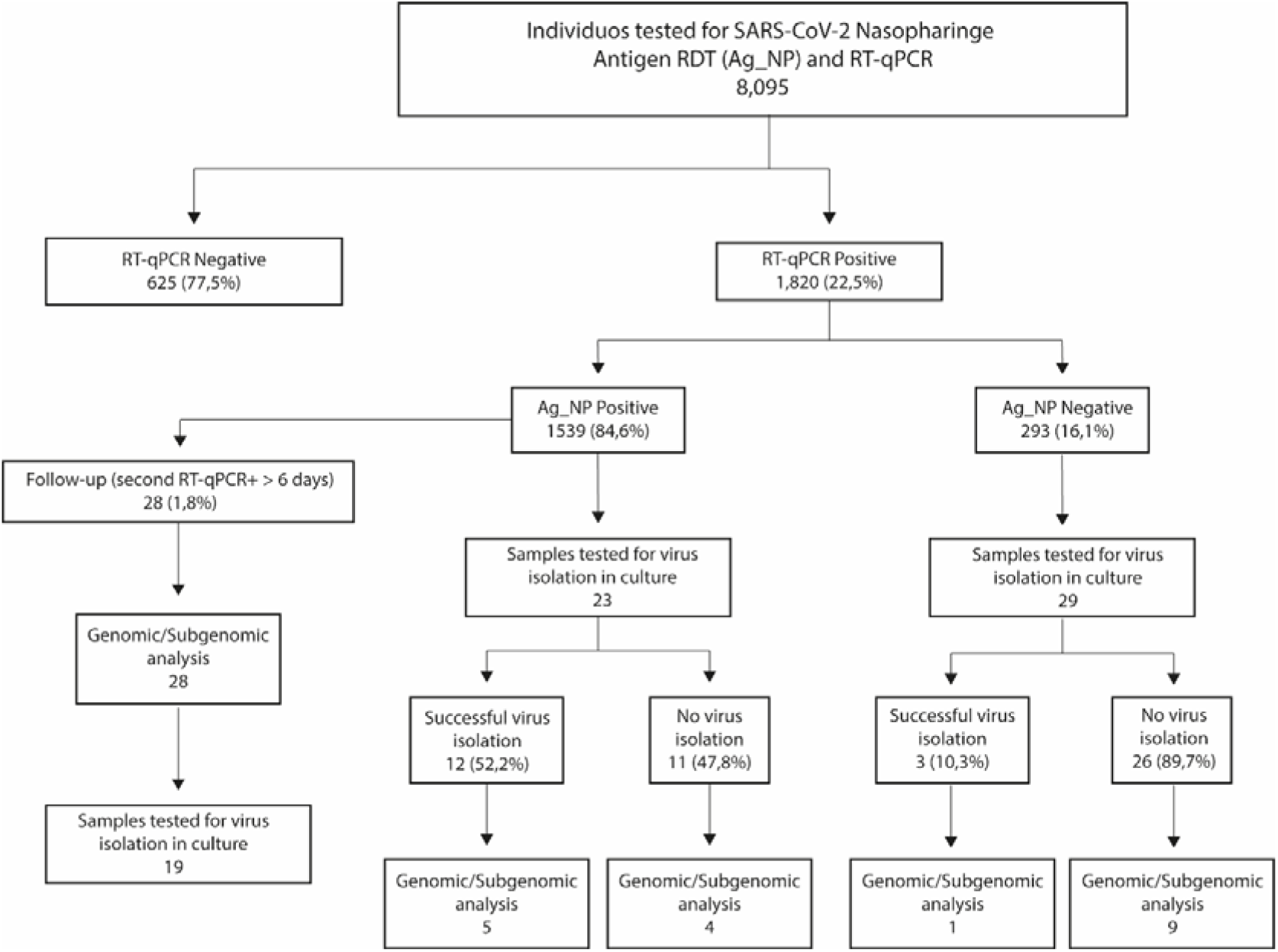
Schematic representation of the UFRJ-CTD cohort during August 20^th^ through September 21^st^ and sample selection. Mildly symptomatic individuals tested for SARS-CoV-2 diagnosis at the Center for COVID-19 Diagnosis of Federal University of Rio de Janeiro from August 2020 to September 2021 account for a total of 8.095 RT-qPCR and antigen RTD exams. Of this total, 1.820 tested positive for SARS-CoV-2 1.539 also being antigen RDT positive (concordant samples) and 293 antigen RDT negative (discordant samples). From this group, 61 samples (23 concordant and 29 discordant) were selected for further studies comprising the cross-sectional dataset while 30 additional patients were selected for the follow-up dataset where the patient has a second samples collected with a minimum of 6 days after diagnosis.

### Cell lines

African green monkey kidney cells (Vero E6, ATCC CRL-1586) and the humanized Vero expressing hAce-2 and hTMPRSS2 (ATCC), referred as Vero-hA/T, were maintained in DMEM (ThermoFisher Scientific) supplemented with 10% fetal bovine serum (FBS; Gibco), 100 U/mL penicillin, and 100 μg/mL of streptomycin (ThermoFisher Scientific). The cells were incubated at 37°C with 5% of CO_2_ and passed every 3-4 days.

### Viral isolation

For viral isolation Vero E6 or Vero-hA/T were plated in six well plates 24 hours before inoculation to achieve 70% of confluence overnight. Cells were infected with 250μL of VTM diluted in DMEM plus 300 U/mL penicillin and 300 μg/mL of streptomycin, with no addition of FBS for 1 h for viral adsorption. After the adsorption period, 10% FBS-supplemented DMEM with 300 U/mL penicillin and 300 μg/mL streptomycin was added to the inoculum and the cells were incubated at 37 °C and 5% CO_2_ for 72 h (passage #1). The supernatants of passage #1 were collected and 250 μL was used to infect fresh cell cultures as previously described, and incubated for another 72h (passage #2), and the remaining supernatant was stored at −80°C for RNA extraction and RT-qPCR for SARS-CoV-2 detection. Passage #2 supernatants were also collected and stored at −80°C for viral titration, RNA extraction and RT-qPCR for SARS-CoV-2 detection. For both cell passages, 100 µL of the collected supernatant was used to perform the Panbio™ COVID-19 Ag assay. For viral titration, 10-fold dilutions of each sample from passage #2 were used to infect Vero E6 cells. After 1 h of virus adsorption, media was replaced by fresh DMEM with 1% FBS, 100u/mL penicillin, 100 µg/mL streptomycin and 1.4% carboxymethylcellulose (Sigma Aldrich) followed by incubation for 4 days at 37 °C with 5% CO_2_. Then, cells were fixed with 10% formaldehyde and stained with 1% crystal violet in 20% methanol for plaque visualization and quantification. Viral titers were expressed as plaque forming units per milliliter (PFU/mL). Positive viral isolation was defined by the visualization of viral plaques after titration of the passage #2 supernatants. If a sample presented a drop of C_t_ value from cell passage #1 to cell passage #2, we performed an additional cell passage (passage #3) as previously described. We considered the sample positive for virus isolation once plaques were visualized in the titration assay. All the experiments were conducted in a BSL-3 laboratory.

### RNA extraction

RNA was extracted from 200 μL of cell culture supernatant from cell passages #1, #2 and #3, when available, using ReliaPrep™ Viral TNA Miniprep System (Promega) according to the manufacturer’s instructions. RNA was eluted in 50 μL and stored at −80°C until RT-qPCR.

### RT-qPCR

For detection of genomic and subgenomic SARS-CoV-2 RNA, we used viral RNA extracted from VTM at the time of diagnosis. Genomic RNA was detected using SARS-CoV-2 (2019-nCoV) CDC qPCR Probe Assay (Integrated DNA Technologies, IA, USA) targeting the SARS-CoV-2 N gene and Brilliant III Ultra-Fast qRT-PCR Master Mix (Agilent). For subgenomic RNA, specific primers for gene N subgenomic product were used together with a probe for N gene along with Brilliant III Ultra-Fast qRT-PCR Master Mix (Agilent) as follows: 0.08 μL of primer forward; 0.08μL of reverse primer; μL of probe; 10μL of 2x master mix; 0.2 μL of 100mM DTT; 0.3 μL of reference dye (dilute 1:500); 1.0 μL of RT/RNase block; 3.0 μL of nuclease-free water and 5 μL of RNA. The cycling used for subgenomic detection was 50°C for 10 minutes; 95°C for 3 minutes; 45 cycles of 95°C for 15 seconds and 53°C for 30 seconds. Sequences of primers and probes are described in Supplementary Figure 3. For detection of genomic RNA from viral cell cultures, we used the AnGene Kit according to the manufacturer instructions. All the reactions were performed using the AriaMX Real-time PCR System (Agilent).

### Sequencing and phylogenetic analysis

Illumina sequencing proceeded using two distinct protocols for library construction, as described in Orf et al (2021) and Moreira et al (2021). Briefly, viral RNA was converted to cDNA with SuperScript IV (ThermoFisher, USA) and whole genome amplification was performed with the ARTIC SARS-CoV-2 V3 primer panel and the Q5 hotstart polymerase (NewEnglandBiolabs, USA). Amplicons were purified and converted into Illumina sequencing libraries with the QIAseq FX library kit (QIAGEN, Germany), following the manufacturer’s protocol. Libraries were quantified with the Qubit dsDNA HS kit and equimolarly pooled, being sequenced on an Illumina MiSeq run with a V3 (600 cycles) cartridge (Illumina, USA).

Raw sequencing data was processed, and reference assemblies were performed using a custom pipeline. Whole genome sequences were then classified using both the pangolin tool v3.1.14 (Pango version 1.2.81) (*26*) and NextClade v1.7.3 (*27*). Afterwards, to further contextualize the novel genome sequences, a maximum likelihood phylogenetic inference was performed with IQ-Tree v2.0.3 (28), under the GTR+F+I+G4 model. The reference dataset was assembled by querying the GISAID EpiCoV database to retrieve the 50 sequences most similar to each of ours, using the Audacity-Instant application (Acknowledgement table available in *Supplementary File 4*). After removing duplicates, the final dataset comprehended 555 genome sequences.

### Western Blotting

For protein analyses, infected-whole cell lysates were collected with RIPA buffer (10 mM Tris-Cl [pH 8.0]; 1 mM EDTA; 0.5 mM EGTA; 1% Triton X-100; 0.1% sodium deoxycholate; 0.1% SDS; 140 mM NaCl) mixed with 0.1% protease inhibitor cocktail (Sigma-Aldrich). Proteins were fractionated by a 4-20% precast gel SDS-PAGE and blotted onto a nitrocellulose membrane (Hybond-ECL, GE Healthcare), using a wet tank transfer system (BioRad). Membranes were probed with primary antibodies anti-Nucleocapsid (cat n°: 26369, Cell Signaling) and Anti-Spike (cat n°: 56996, Cell Signaling). The secondary antibodies used were the horseradish peroxidase HRP-conjugated Anti-Rabbit (KPL). SuperSignal West Pico Chemiluminescent Substrate (Thermo Scientific) was used as a substrate for the horseradish peroxidase (HRP) chemiluminescent reaction.

### Serologic assay

Microtitre plates (Immulon 2 HB) were coated with and trimeric spike protein at 4μg/mL or nucleocapsid protein at 1 ug/mL in 100 mM sodium carbonate-bicarbonate buffer (pH 9.6) and incubated overnight at 4°C. Excess protein was removed by washing five times with PBS + 0,05% Tween-20 (PBST, Sigma) and unbound sites blocked using 5% BSA in PBST. Samples were added at 1/50 dilution in PBST + 2% BSA, followed by 1 hour at 37°C. The plates were washed five times with PBST, and polyclonal anti-human IgG antibody conjugated to HRP (Promega) added for one hour at room temperature. Plates were washed five times with PBST followed by addition of the chromogenic substrate TMB (Sigma) for 10 minutes and reaction stopped with 1N sulphuric acid. Absorbance was read at 450 nm with an ELISA microplate reader (Biochrom Asys). A positive control specimen (from a known COVID-19 patient) in simplicate and a negative control (pre-epidemic plasma sample) in triplicate were added to every assay plate for validation and cut-off determination. Results were expressed as a reaction of sample optical density value divided by assay cutoff.

### Statistical analysis

The data from the cohort was acquired using the KoBoCollect online/offline web-based form (Available at: https://www.kobotoolbox.org). The data was extracted as a dataset based on XLSForms and merged with the laboratory data. Differences between concordant and discordant groups were assessed by Mann-Whitney non-parametric U-test. Multivariate model based on logistic regression was applied for multivariate analysis. GraphPad Prism version 9.2.0 (GraphPad Software, San Diego, California USA), JASP Version 0.16 (JASP Team, 2021). [Computer software]), and R (R Core Team 2021. *R: A Language and Environment for Statistical Computing*, Vienna, Austria. Available at: https://www.R-project.org/) were used. A *p* value < 0.05 was considered significant.

## Results

From August-2020 to September-2021, a total of 8,095 individuals were simultaneously tested for SARS-CoV-2 by RT-qPCR and Panbio™ Antigen RDT tests (Figure 1) at the center for COVID-19 diagnosis of the Federal University of Rio de Janeiro. While 1,820 were positive for RT-qPCR (22.5%), 1,539 (20.34%) were positive for the Ag-RDT (Figure 1), yielding 84.6% of concordant (RT-qPCR+/Ag-RDT+) and 16.1% of discordant (RT-qPCR+/Ag-RDT-) samples (Figure 1). The total number of samples tested per month varied from 159 (Jun 2021) to 1208 (Nov 2020), with a positivity rate/month of 15% (Sep 2021) to 36% (Nov 2021). Regardless of the transmission rates, the percentage of discordant samples did not vary substantially over time (Figure 2).

**Figure 2.**
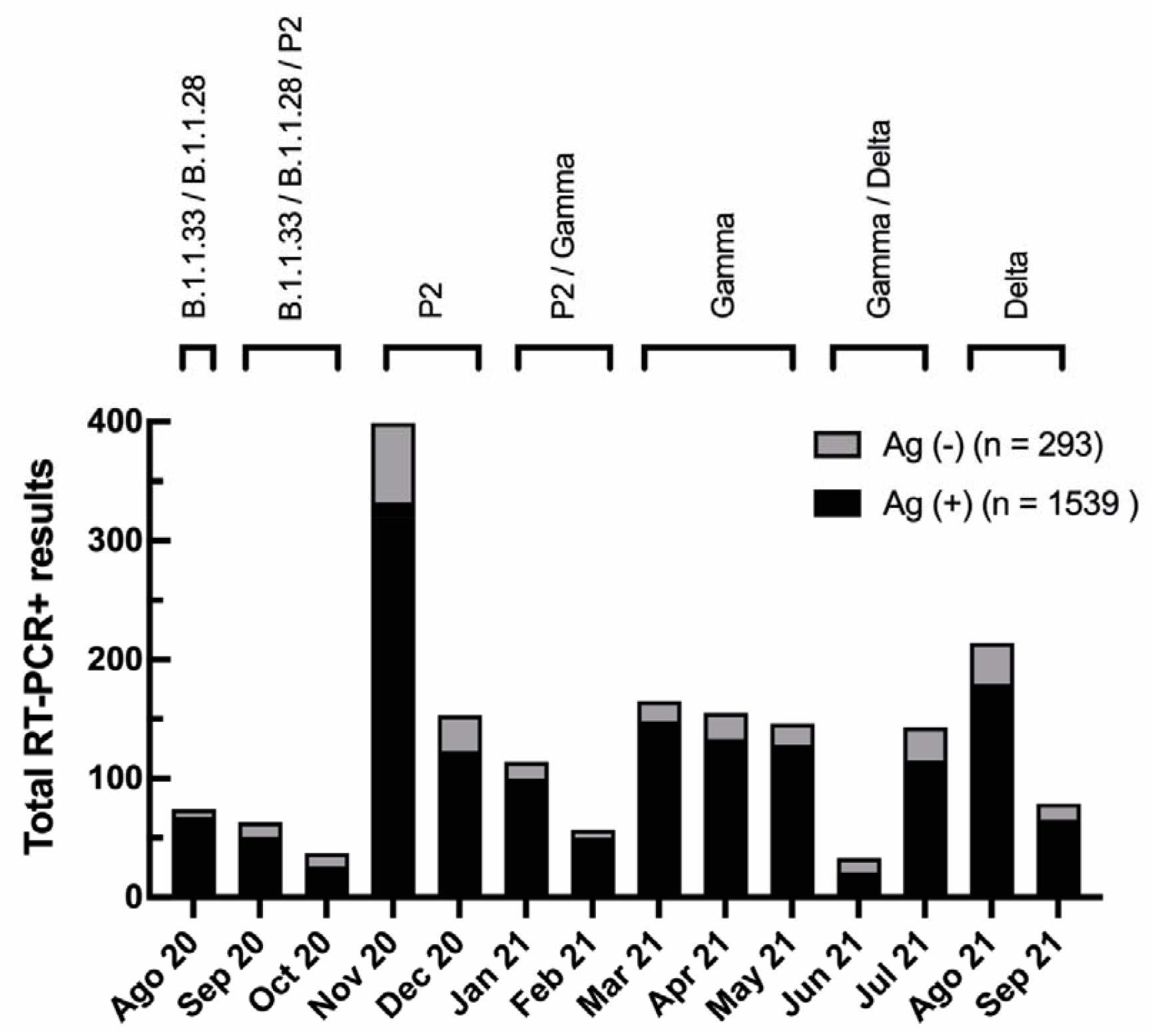
Evaluation of Panbio™ COVID-19 Antigen RDT test showed low false negative occurrence with no variation during the time of the study. Total number of RT-PCR positive results per month in the studied population. In black, total number of RT-PCR positive individuals with a negative Antigen [Ag (-)] RDT result (discordant group). In gray, total number of with RT-PCR positive individuals with a positive Antigen [Ag (+)] RDT result (concordant group). At the top, prevalent viral variants in Rio de Janeiro state during the study, according to data from the Vigilance Network of the State of Rio de Janeiro (blob:http://www.corona-omica.rj.lncc.br/4efa46e4-9323-452e-b3b2-c8014526a9ad).

Different SARS-CoV-2 variants circulated in Rio de Janeiro during this period (Figure 2), suggesting that Ag-RDT sensitivity was not impacted by these viral variants and neither with by the beginning of the vaccination program in January 2021 either.

Using a logistic regression analysis model, we demonstrated that RT-qPCR C_t_ value, DSSO, and IgG anti-S significantly contribute to the probability of detecting a positive Ag-RDT. The RT-qPCR C_t_ value had the highest impact (Supplementary Figure 1). The month of diagnosis could reflect a potential difference in the level of viral circulation, the prevalence of different variants over time, and the percentage of vaccinated individuals. However, it did not affect the probability of having a positive Ag-RDT result, suggesting that these factors did not alter Ag-RDT sensitivity (Supplementary Figure 2).

Ag-RDT negative samples had higher C_t_ values (Figure 3A) than Ag-RDT positive ones, with a positive correlation between C_t_ values and DSSO among the RT-qPCR positive samples (Figure 3B). This correlation was maintained for Ag-RDT positive (concordant) (Figure 3C) but not for Ag-RDT negative (discordant) samples (Figure 3D), probably due to their higher C_t_ values.

**Figure 3.**
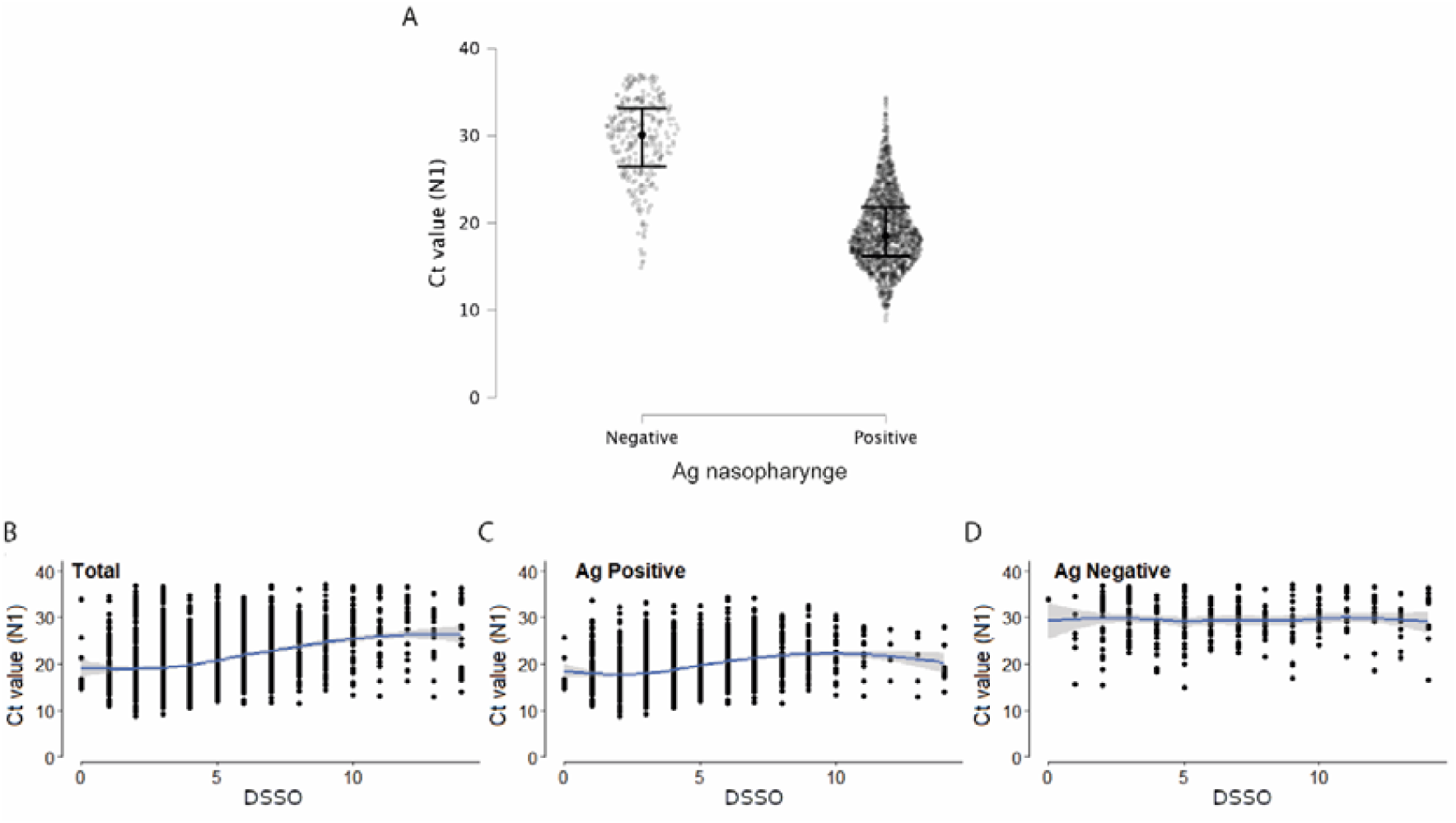
Discordant samples (RT-PCR+/Ag-RDT-) showed higher Ct values than concordant ones (RT-PCR+/Ag-RDT+). *A:* Ct values (N1 target) of SARS-CoV-2 RT-PCR in patients with a positive (n = 1,539) or negative (n = 293) nasopharyngeal antigen (Ag) rapid diagnostic test (RDT) result tested from Aug 2020 to Sep 2021. *B - D:* Correlation between Ct value (N1 target) of SARS-CoV-2 RT-PCR and days since symptom onset (DSSO) when samples were collected in total patients (B), and among patients with a positive or negative Ag-RDT result (C and D, respectively).

To further understand what influenced the sensitivity of the Ag-RDT when compared to the RT-qPCR assay, a total of 68 samples were selected (23 concordant and 29 discordant). We analyzed the C_t_ value at the time of diagnosis for all samples in both study groups. Discordant samples notedly had higher C_t_ values (14-28, Median = 21) when compared to concordant samples (20-38, Median = 28) (Figure 4A), as demonstrated above for the total number of samples (Figure 3A).

**Figure 4:**
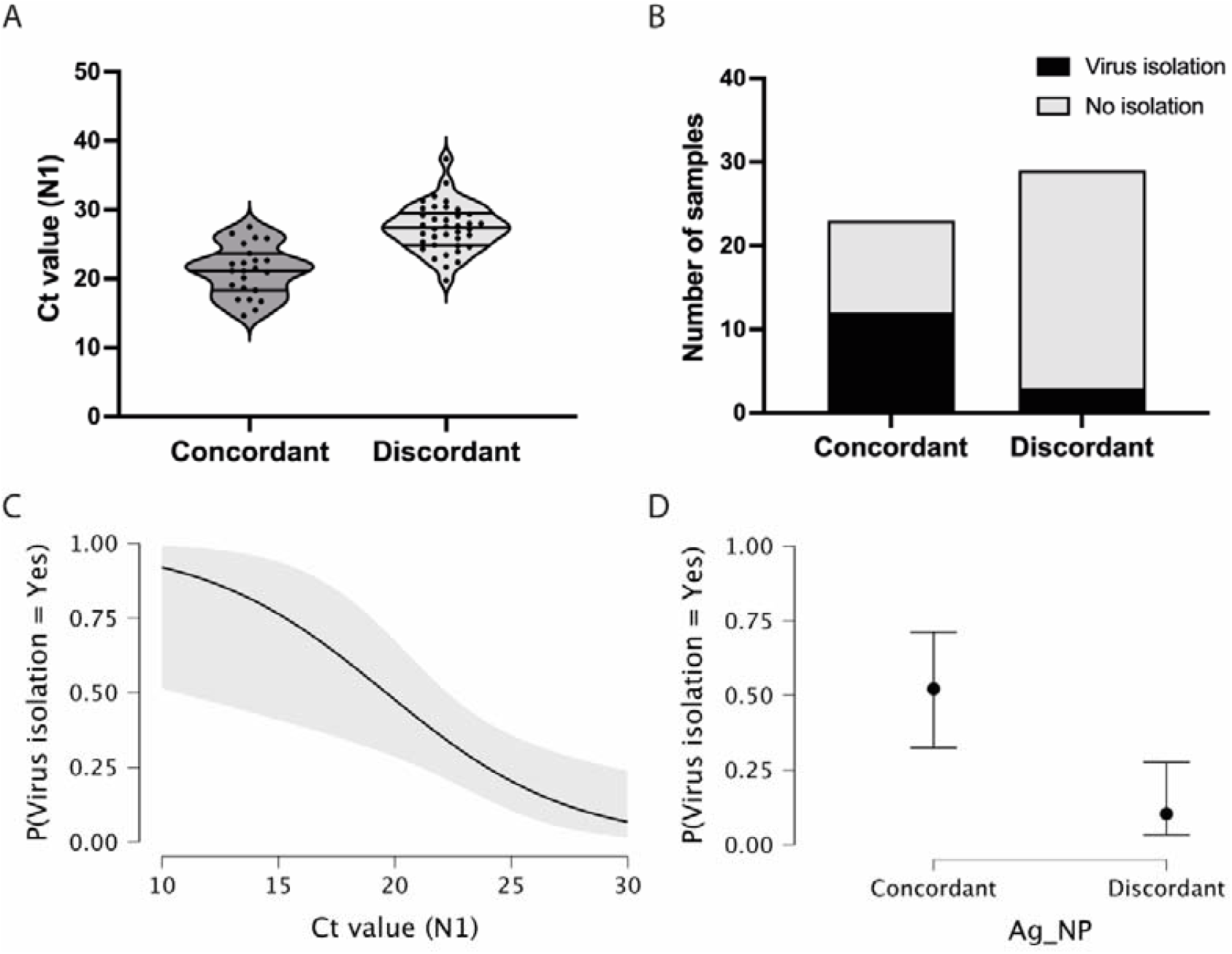
Antigen discordant samples have higher C_t_ and lower viral isolation rate compared to discordant samples. *A*: Violin plot showing RT-PCR C_t_ values (N1 target gene) in antigen concordant (RT-PCR+/Ag-RDT+, n = 23) and discordant (RT-PCR+/Ag-RDT-, n = 29) samples used for virus isolation experiments, lines represent the median and 25%-75% quartiles. *B*: Virus isolation success in total number of antigen concordant (n = 23) and discordant (n = 29) samples. *C-D*: Viral isolation probability considering RT-PCR Ct values (N1 target gene) and nasopharyngeal antigen rapid test (Ag-NP) result as isolated covariates, respectively.

We evaluated the presence of viable infectious viruses in concordant and discordant samples after two consecutive cell passages. Positive virus isolation was only confirmed after virus titration using plaque assay. From the 23 concordant samples, 12 samples were positive for virus isolation (rate = 52.17%). In contrast, for the 29 discordant samples, only 3 were positive for virus isolation (rate = 10.34%) (Figure 4B). These percentages indicate a significant difference in the isolation success between Ag-/RT-PCR+ and Ag+/RT-PCR+ groups (*p* < 0.00001, CI 0.05). Analyzing viral genomic RNA levels in the supernatants from first passage cultures, 7 out of 23 concordant samples (30.43%) and only 1 out of 29 discordant samples (3.45%) predicted virus isolation already after the first passage (C_t_ levels < 25 - Supplementary Table 1), suggesting a relatively higher abundance of infectious viruses present in concordant samples. When the sample C_t_ is considered as an indicator of virus isolation success, the higher probability of virus isolation significantly occurred for both concordant and discordant samples in the low C_t_ range (10-20). This probability dropped to less than 25% with C_t_ values greater than 26 (Figure 4C). When analyzing discordant and concordant samples separately for virus isolation success in the same C_t_ range, the probability of virus isolation was below 10% for the former and above 50% for the latter (*p* = 0.002).

The absence of viral particles in nasopharynges still positive for total viral RNA could be due in major part to the presence of viral subgenomic RNA (sgRNA). To verify the influence of sgRNA on the C_t_ value, we performed in parallel target-specific RT-qPCR for genomic and N-sgRNA in a few concordant and discordant samples (Supplementary Figure 3A). We observed that discordant samples presented higher C_t_ values for both genomic and N-sgRNA than concordant samples (Supplementary Figure 3B). Since sgRNA is a marker of viral replication, this data suggests a lower level of replication in antigen discordant samples, which correlates to a lower proportion of these samples harboring infectious viruses. Importantly, from this data, we showed that N-sgRNA proportionally contributed 20% of the total amount of the viral RNA present in both concordant and discordant nasopharyngeal samples (Supplementary Figures 3C). These results suggest that RT-qPCR positive, but antigen negative nasopharyngeal samples, lack competent/infectious viral particles in part due to a lower level of viral replication, thus having a lower impact on the spread of SARS-CoV-2.

Next, we used the VTM and isolated viruses to characterize the full-length genomic viral RNA from a few concordant and discordant samples. We recovered intact full-length genomes from 8 discordant (4 VTM and 4 virus isolates) and 14 concordant (7 VTM and 7 virus isolates) samples. The phylogenetic reconstruction demonstrated the clustering of these samples in 3 distinct clades: B.1.1.33; Brazilian P.2 (Zeta); and P.1 (Gamma), in agreement with the period samples were collected (Figure 5A). Distinct clustering was not observed when we analyzed discordant versus concordant samples, suggesting that no distinct genome characteristic would account for the discordant status. Analyzing specifically the N ORF, no major mutations or insertion/deletions were present in the discordant samples, excluding the possibility that gross genomic differences could compromise the recognition of the N protein by Ag-RDT (Figure 5B), and the nucleocapsid mutation profiles were largely identical for both groups (Figure 5B). Sequences recovered from both VTM and isolated virus from discordant sample 37376 were identical.

**Figure 5:**
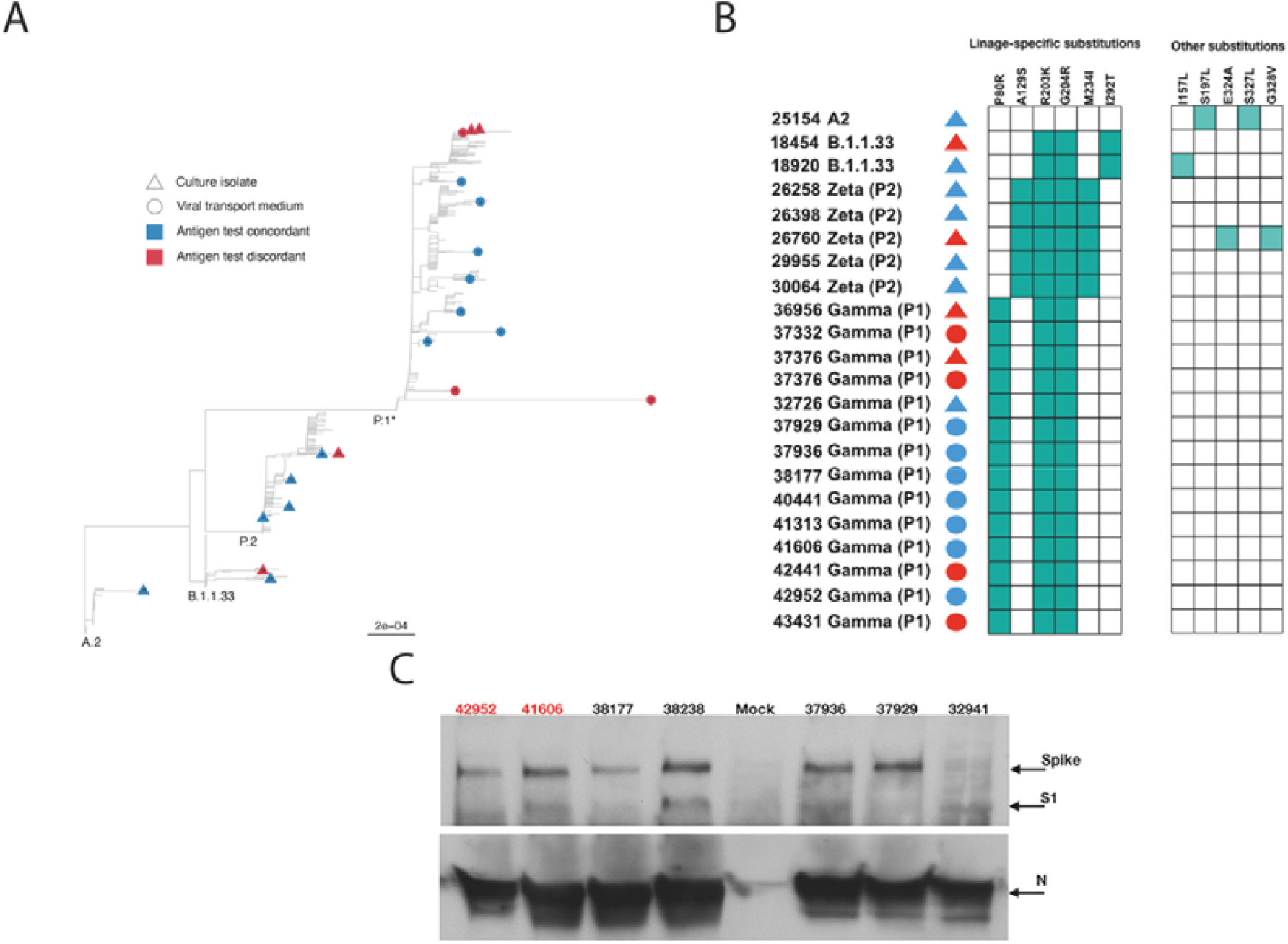
Phylogenetic and protein analysis indicate no major differences between concordant and discordant samples. *A:* Phylogenetic tree of VTM and isolated viral samples demonstrated that concordant and discordant samples do not form specific clusters and grouped only according to the viral lineage. *B:* Schematic representation of amino acid differences in N gene between sequenced VTM or viral isolated concordant and discordant samples. *C:* Detection of SARS-CoV-2 Spike, S1, and Nucleocapsid (N) proteins by Western blotting in lysates from Vero E6 cells infected with concordant and discordant samples. Uninfected cells were used as mock. Antigen discordant samples (42952 and 41606) in red and antigen concordant samples (38177, 38238, 37936, 37929, 32941) in black.

Both S and N proteins were readily detected from infected cell lysates by Western blotting and no major difference in S and N amounts was observed (Figure 5B). The anti-N antibody equally detected the N protein from viruses recovered from both discordant and concordant samples. These data demonstrate that although discordant samples failed to be detected by the antibodies in the Ag-RDT, it was probably due to the small amount of intact viral particles in the nasopharyngeal samples rather than a lack of reactivity of kit capture monoclonal antibody with the N protein.

Next, we hypothesized that patients with a prolonged infection present reduced viral shedding thus accounting for the discordance between the Ag-RDT and RT-qPCR results. To evaluate the impact of the time of infection in antigen detection, we studied a cohort of 30 patients longitudinally, totaling 41 follow-up specimens, that were collected from January to September 2021, where the first patient sample was mandatorily concordant (RT-qPCR+/Ag-RDT+) and the patient had at least one more sample collected after the first diagnosis. All sample information is summarized in Supplementary Table 2. The minimal time interval between the first (day 0) and the following sample collection varied from 6 to 32 days. On day 0, the sample C_t_ ranged from 12-26 followed by an overall increase in the C_t_ value for viral RNA over time (Table 1 and Figure 6). We observed that N-sgRNA values also increased and antigen positive results decreased as time passed since symptom onset (Table 1). Therefore, a prolonged SARS-CoV-2 infection increases the likelihood of a negative Ag-RDT result without impacting detection by RT-qPCR. Importantly, out of 5 follow-up samples that were still positive for the Antigen RDT, 3 were collected early after symptom onset (6-8 days). The VTM of 70 samples was used to infect Vero-hA/T cells, followed by plaque titration of passage #2 supernatants. Among the 29 samples from day 0, 21 produced infectious viruses (rate = 74.41%). Among the 36 discordant follow-up samples viruses were isolated from 5 (rate = 13.89%) showing a clear drop in virus isolation with DSSO increase (Table 1 and Supplementary Table 2). This data shows that most upper respiratory tract discordant samples do not harbor infectious viruses and consequently would be less transmissible.

**Table 1:**
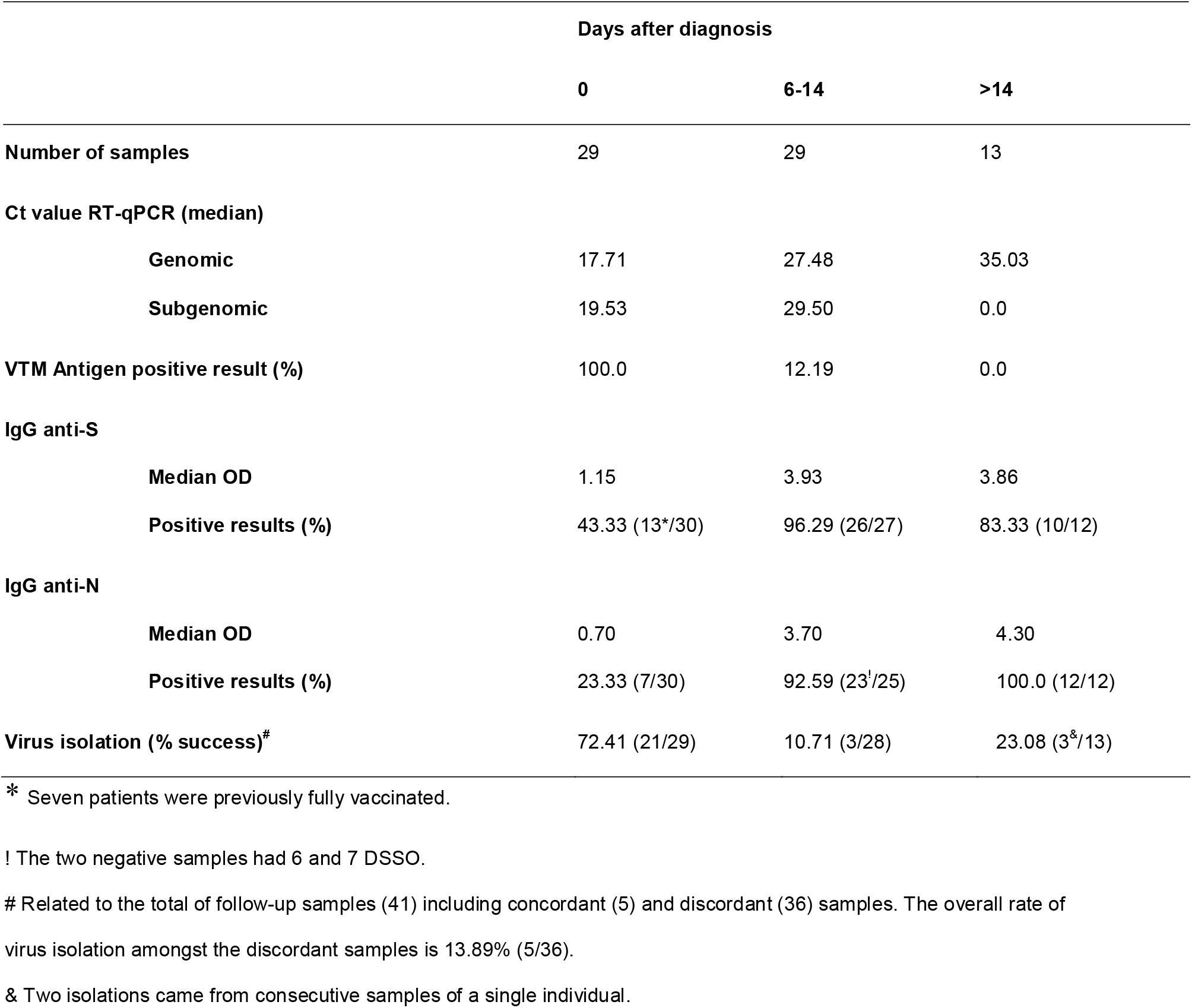
Summarized data from follow-up samples.

**Figure 6:**
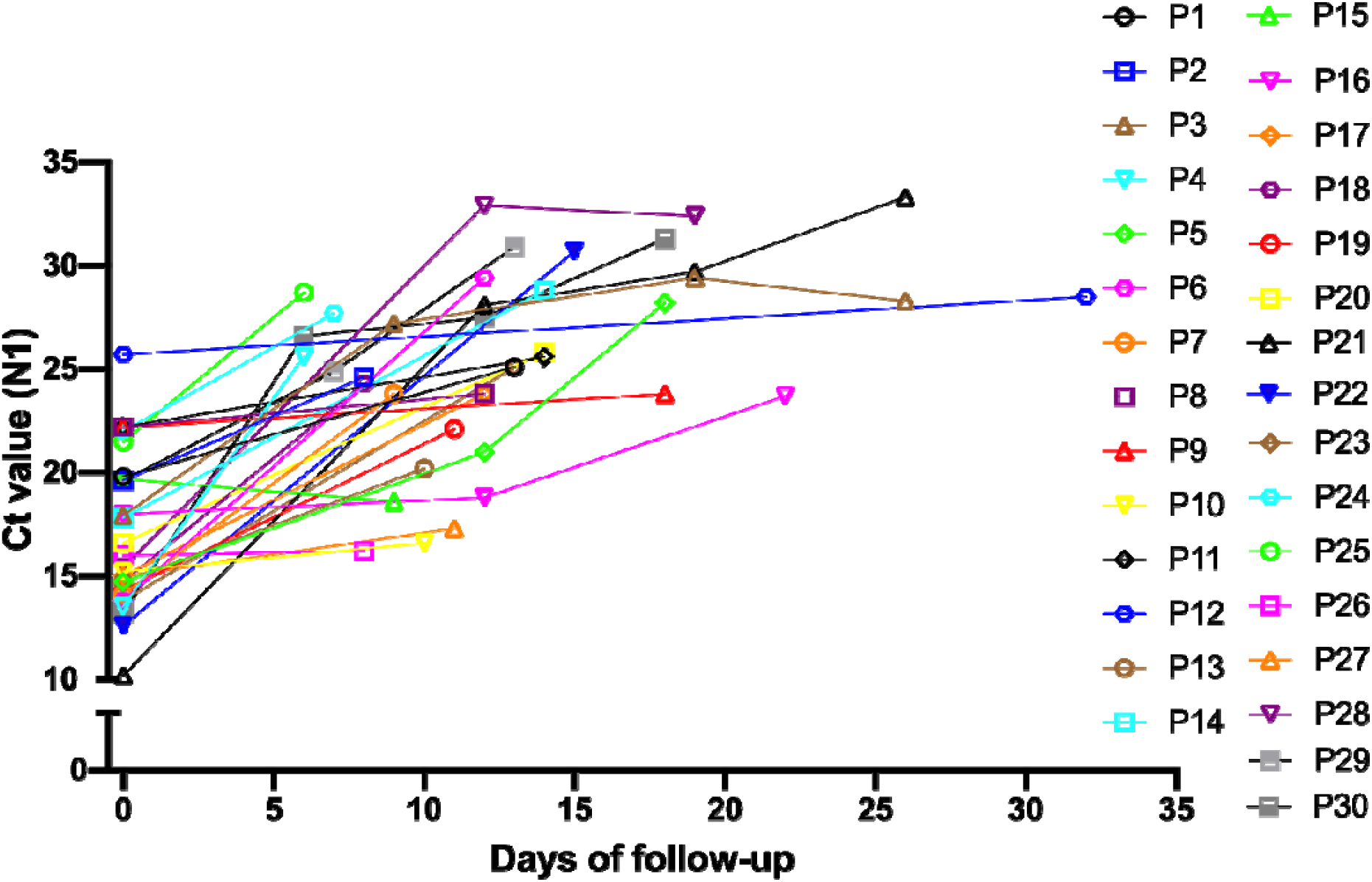
C_t_ values increase with prolonged infectious periods in follow-up samples. Graphical representation of the C_t_ value from the 30 patients that comprised the follow-up dataset at the first sample collection and the following samples from these individuals during the follow-up.

We demonstrated that a prolonged infection period decreases the likelihood of viral particle detection by the Panbio™ Ag-RDT which in our data set is overall correlated with lower viral loads in nasopharyngeal samples as well as a lower level of viral replication/infectivity. However, a few discordant samples had viral loads in the range of 18-25 which predicted a substantial amount of viral replication and the presence of infectious viruses. Then, we asked whether the establishment of the anti-SARS-CoV-2 antibody response in those individuals would contribute to a decrease in detection by the Ag-RDT. The presence of serum IgG anti-S and anti-N antibodies was measured in initial and followed-up samples. We observed that 92.59% and 100% of samples collected 6 to 14, and >14 days after diagnosis, respectively, had anti-N IgG. 96.29% and 83.33% of samples collected 6 to 14, and >14 days after diagnosis, respectively, had an anti-S IgG response (Table 1). Amongst initial diagnostic specimens, 23.33% already had anti-N IgG and 43.33% had anti-S IgG responses (Table 1). It is noteworthy that 15.78% of the diagnosed patients had been previously fully vaccinated. Conversely, 87.88% (36/41) of the individuals had a negative Ag-RDT in the followed-up sample post-seroconversion for both anti-S and anti-N IgG. Taken together, these data suggest that a prolonged SARS-CoV-2 infection predicts a lower viral load and virus replication in the nasopharynx which can be associated with the presence of circulating anti-N and anti-S immunoglobulins and the decreased detection by Ag-RDT.

## Discussion

Since the COVID-19 pandemic remains a public health challenge even after the rapid development and increased access to available vaccines, it is important to ensure testing capacity with fast and reliable methods to guide patient isolation to curb virus transmission.

Our study cohort was composed of two datasets: a cross-sectional, where the Panbio™ COVID-19 Ag test was only performed in symptomatic patients, and a follow-up dataset in which at least two samples were collected from the same patient for the Ag-RDT with a minimum of 6 days between sample collection. For both datasets, the viral isolation rate was higher for the concordant samples (56.52 - 66.7%) when compared to the discordant ones (10.53 - 10%) even for the same C_t_ range (14-29 versus 18-29). For discordant samples, viruses could not be cultured when C_t_ values were higher than 30, confirming previous reports of low or absence of infectious viruses at this C_t_ range (*29,30*). We observed that the virus isolation rate is not altered by different viral variants or sample collection. In different epidemiological weeks, RT-qPCR results indicated higher C_t_s for discordant samples for both genomic and sgRNA compared to concordant samples, and the relationship between genomic and sgRNA demonstrated that sgRNA levels do not impact the C_t_ of diagnosis or the isolation success in both groups.

The virus isolation criteria used in this article relies on a plaque titration assay for determination of positive or negative sample isolation. Thus, only samples harboring replication competent viruses were considered, which is a proxy for specimen infectivity, provides a more reliable measurement of viral viability/infectiousness than RT-qPCR. Our results demonstrated that discordant samples present lower levels of viral replication meaning that a patient with a negative Ag-RDT result has a reduced risk of transmission even when presenting as RT-qPCR positive, indicating that Ag-RDT has high epidemiological relevance.

Then, the use of Ag-RDTs could be a powerful tool for detecting infected individuals with higher potential for transmission. Its specificity and sensitivity correlate with time from symptom onset and the presence of viral infectious particles. Data on the use of antigen tests in large cohorts like universities, healthcare institutions or even in music events as a SARS-CoV-2 screening method demonstrated that sensitivity depends on the target population where the test is used. In general population screening, antigen tests had a reported sensitivity from 41.2 to 74.4% in different countries (*31,32,33*), and in emergency rooms and nursing homes the sensitivity varied from 50% to 92% even for asymptomatic individuals (*34,35*). Revollo and colleagues demonstrated the utility of Ag-RDTs for same day screening in mass gathering events, where no infected individuals were detected in follow-up tests 8 days after the event, indicating that the antigen test can detect potential transmitters (*36*). Despite the variation in sensitivity, a recent study using mathematical modeling showed that Ag-RDTs could be more efficient than RT-qPCR in reducing SARS-CoV-2 transmission by up to 85% in a nursing home facility (*37*). Another study predicts that Ag-RDTs could prevent infections better than RT-qPCR in a workplace (*38*).

As vaccination campaigns progress around the world, high vaccine coverage in several regions have been achieved (*1*). Nevertheless, places like continental Europe and the US are experiencing new waves of infection with thousands of new cases daily. Although vaccines present a greater than 90% protection against severe disease (*39,40*) their effectiveness has decreased substantially (*41,42*). One of the factors related to the decrease in effectiveness is the emergence of new SARS-CoV-2 variants. An effectiveness reduction from 91.78% to 79.87% was reported in New York city during May to July 2021 as well as the detection of breakthrough cases among fully vaccinated individuals infected with the Delta variant (*43,44*).

New variants are mainly classified according to mutations in Spike protein, however since mutations also accumulate in the N gene, they could impact the results of Ag-RDTs targeting N. The samples used in this study were collected throughout waves of different variants that circulated in Rio de Janeiro from August-2020 to September-2021, including the Zeta, Gamma and Delta lineages. Our results agree with others that did not observe any impact in Ag detection of these variants (*45,46*). The presence of substitutions T205I and D399N in the N ORF were linked to a failure in Ag-RDT detection (*47*). We observed few mutations in concordant and discordant samples, but none impacted on antigen recognition. The N and S proteins from viruses isolated from discordant samples were readily detected by Western-blot emphasizing the fact that when present in adequate amounts and/or conditions, no difference in antigen-antibody recognition for all samples will exist.

It was clear that a specific and still unrecognized condition of the discordant sample would account for the Ag-RDT negative result and lower probability of virus isolation even with the presence of high amounts of intact full-length viral RNA. The characterization of samples in the follow-up dataset narrowed down this specific condition to a phenomenon related to time past from initial symptoms, which could be for instance patient’s seroconversion, although the presence of circulating serum anti-N and anti-S IgG does not necessarily correlate with mucosal immunoglobulins. Thus, the establishment of an upper respiratory tract environment over time of infection could induce virus aggregation/reduced stability leading to low viral shedding and missed antigen capture in Ag-RDT.

Recent data demonstrated a higher rate of disease progression in individuals with no detectable antibody and, like our results indicate, the disappearance of antigen correlated with the rise of antibodies (*48*). These data suggest that a positive Ag-RDT indicates active disease, and a longer time for viral RNA clearance than antigen.

Considering the variable Ag-RDT sensitivity, its implementation for mass rapid testing is valuable to identify highly infectious individuals in a community, as most Ag-RDT negative samples probably will not harbor infectious viruses. Therefore, it may assist health authorities to control the virus spread while avoiding unnecessary individual isolation precautions.

## Conclusions

This study found a clear association between a positive Ag-RDT result and the presence of infectious virus in COVID-19 nasopharyngeal exudates. A positive Ag-RDT result was also correlated to a C_t_ <30 in our RT-qPCR test, accompanied by the absence of measurable IgG against the S and N viral proteins and less than 7 days after symptom onset. Our data also supports the proposal to identify patients using Ag-RDT testing in the general population to substantially reduce the number of infected individuals with higher risk of viral transmission while avoiding unnecessary individual isolation.

Massive scaling of testing with Ag-RDT and patient isolation together with an extensive vaccination campaign should reduce the prevalence of SARS-CoV-2 infection and, consequently, the number of severe cases needing hospitalization, leading to a lower COVID-19 death toll.

## Data Availability

All data produced in the present study are available upon reasonable request to the authors.

## Disclaimers

## Supplementary Figures

**Supplementary Figure 1.**
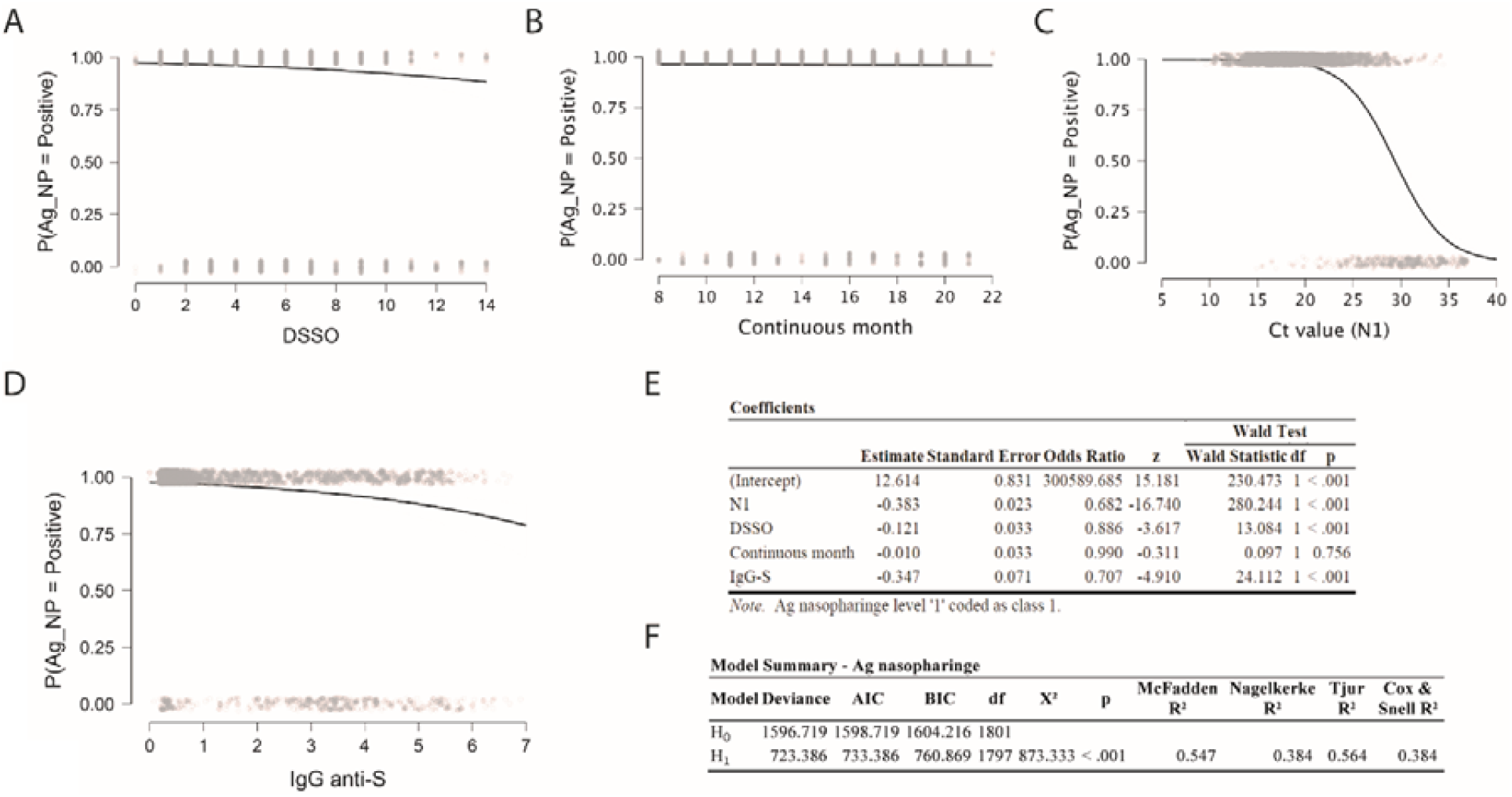
SARS-CoV-2 RT-PCR Ct value and IgG anti-S levels highly influence the chance of having a positive nasopharyngeal antigen test result. Logistic regression showing the probability of having a positive nasopharyngeal antigen (Ag_NP) result according to: (A) SARS-CoV-2 RT-PCR Ct value (N1 target); (B) IgG anti-S level; (C) days since symptom onset; and (D) month of sample collection, from Ago 2020 to Sep 2021 (continuous month 8-21). (E and F) model summary and coefficients of logistic regression showing the probability of having a positive nasopharyngeal antigen (Ag_NP) result according to SARS-CoV-2 RT-PCR Ct value (N1 target); IgG anti-S level; days since symptom onset; and month of sample collection, from Aug 2020 to Sep 2021 (continuous month 8-21).

**Supplementary Figure 2.**
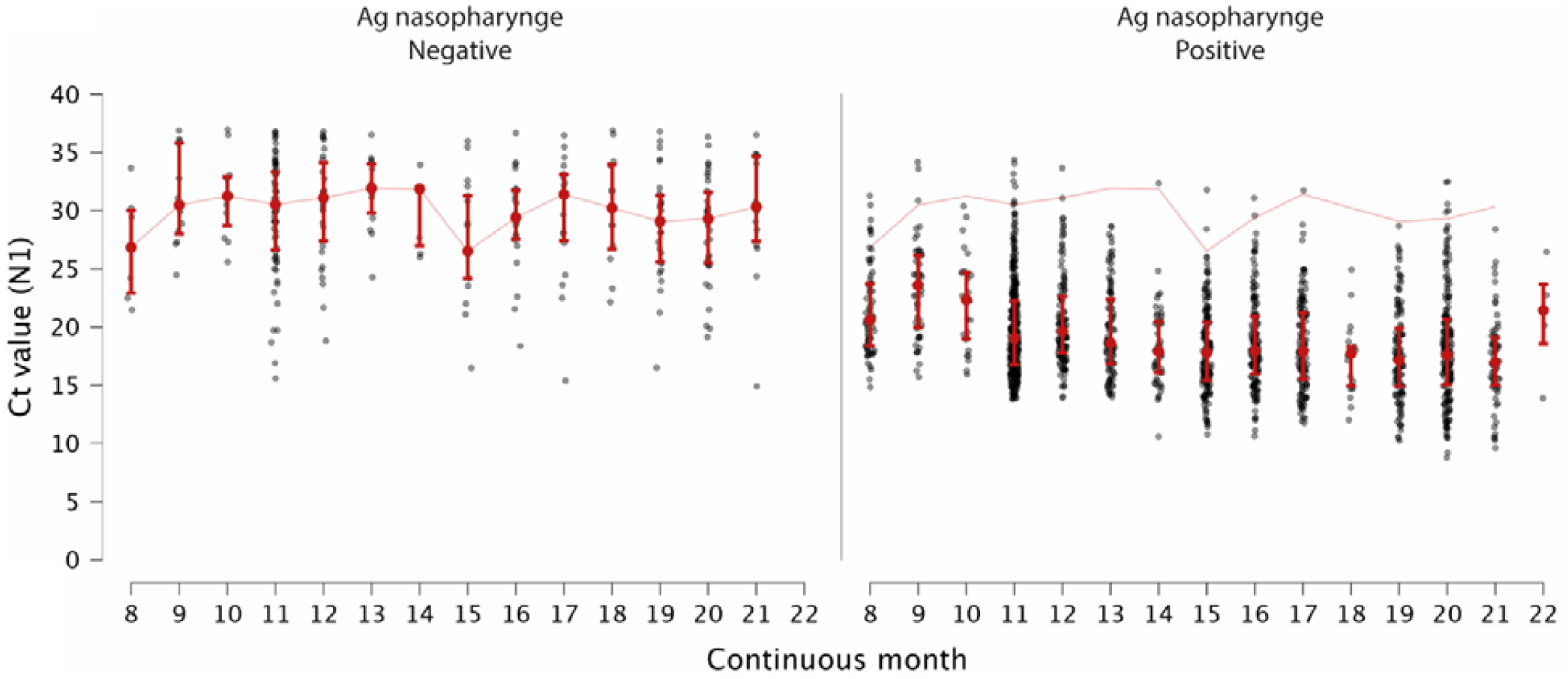
Continuous analysis of C_t_ values from samples RT-qPCR positive results demonstrated that samples with antigen RDT positive results had lower Ct values than antigen RDT negative ones. Ct values (N1 target) of SARS-CoV-2 RT-qPCR in patients with positive or negative Ag-RDT results by month, from August 2020 to September 2021 (continuous month 8-21).

**Supplementary Figure 3.**
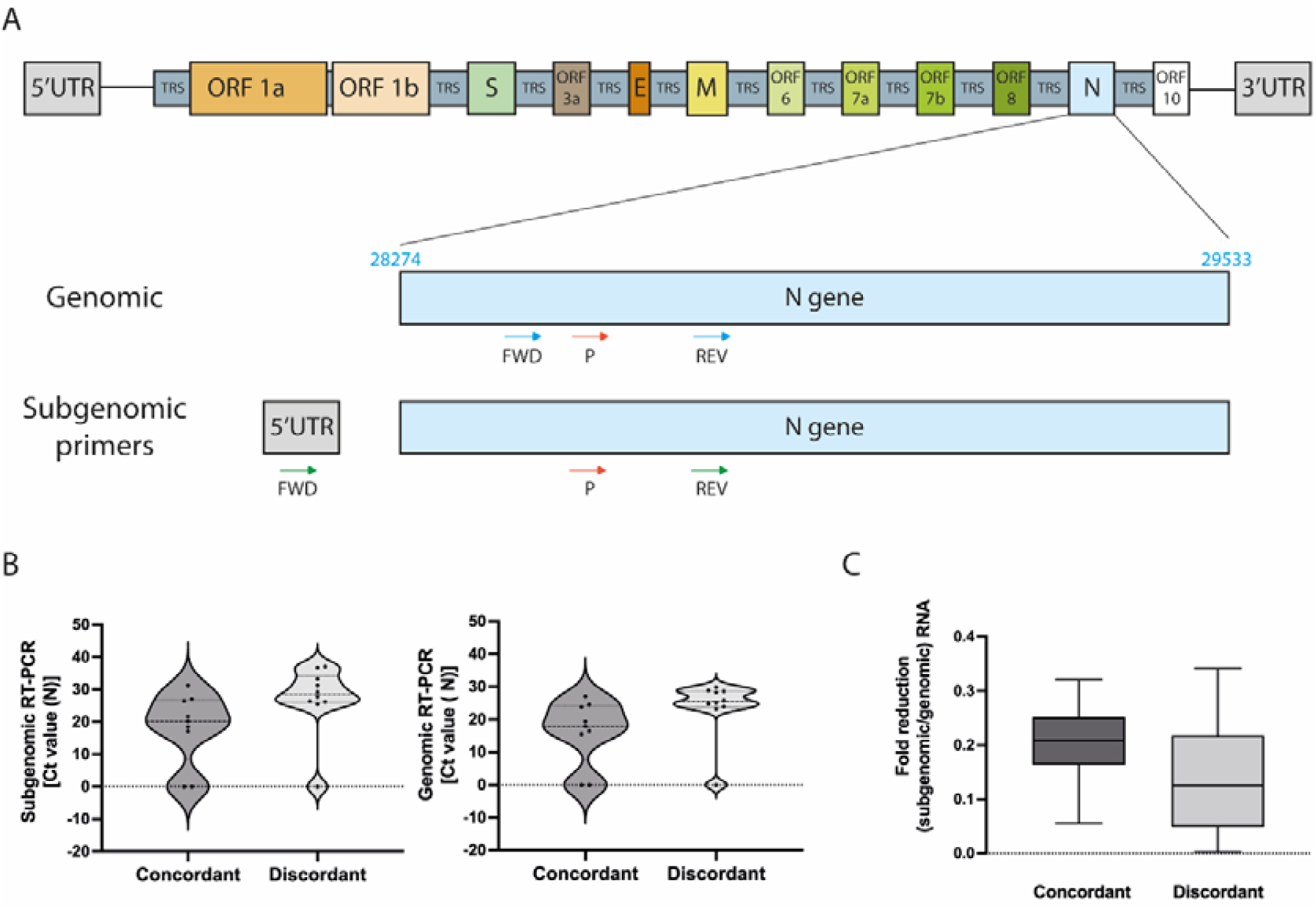
Antigen concordant and discordant samples had similar genomic to subgenomic RNA ratio. *A:* Schematic representation of primers and probes used for RT-qPCR detection of genomic and subgenomic RNA in VTM samples. Primer sequences used for genomic detection are N_sarbeco_F - CACATTGGCACCCGCAATC and N_sarbeco_R - GAGGAACGAGAAGAGGCTTG. For subgenomic detection, primers sequences are: FWSGRNAN - CGATCTCTTGATCTGTTCTCTAAACGAACAAATTAAAATG and N_sarbeco_R - GAGGAACGAGAAGAGGCTTG. For both reactions, the probe used was N_sarbeco_P - FAM-ACTTCCTCAAGGAACAACATTGCCA-BBQ. *B:* Median Ct value from genomic RNA (N gene) from antigen positive and negative VTM samples. *C:* Fold reduction calculated based on the delta Ct of genomic and subgenomic ratio.

